# Predictors of outpatient hysteroscopy pain: A role for retrospective cervical screening pain estimates? A service evaluation

**DOI:** 10.1101/2025.05.29.25327068

**Authors:** Richard Harrison, Nico Biagi, William Kuteesa

## Abstract

Outpatient hysteroscopy(OPH) is a key diagnostic approach in gynaecology, valued for its cost efficiency and avoidance of general anaesthesia. However, severe pain affects 15–34.8% of patients, compromising procedure success, as well as patient well-being and satisfaction. Despite this, predictors of pain remain underexplored. This service evaluation examined data from 804 patients undergoing OPH. Pre-procedure variables, including prior procedural pain (e.g. cervical screening), childbirth history and clinical details, were assessed using stepwise logistic regression to predict patient-reported pain levels.

Retrospective pain ratings from previous cervical screenings emerged as the strongest predictors of OPH pain. Patients who recalled moderate or severe pain during cervical screening were significantly more likely to report severe pain during OPH(p < 0.001). Multiparity was linked to reduced pain perception, although interestingly, this doesn’t appear to be associated with vaginal vs Caesarean delivery. Diagnostic hysteroscopy is likely to be more painful than polypectomy which is likely due to the inclusion of endometrial biopsy when undertaking a diagnosis. Similarly, IUD removal, which requires no biopsy, is less likely to elicit pain than diagnostic investigation of pre or post-menopausal bleeding, which does.

These findings highlight the potential of pre-procedural pain history, particularly from cervical screening, as an easily applied predictor for OPH pain. Overall, this approach could be valuable in identifying patients at risk of a painful procedure. Ultimately, this approach could be optimised to facilitate predictive stratification of patients to sedation, which could enhance success rates in OPH and protect patient well-being. This study introduces a range of presurgical risk factors, including cervical screening pain history as a novel, domain-specific predictor. This offers an opportunity to personalize patient care, reduce the burden of severe pain, and improve procedural outcomes in OPH.

**Plain Language Summary:** Outpatient hysteroscopy (OPH) is a widely used gynaecological procedure for examining the uterine cavity. As an outpatient procedure, OPH is cost-effective and aligns with healthcare policies aimed at reducing hospital admissions. However, it is frequently associated with significant pain, with up to 85% of patients reporting pain during the procedure. Identifying patients at risk of severe pain is critical to improving the patient experience and reducing procedure failure rates.

This service evaluation analysed data from 804 women undergoing OPH at a UK hospital to identify factors that predict pain levels. Analysis revealed that women who experienced prior painful cervical screening were more likely to report higher pain during OPH. Other predictors included the type of procedure or the patient’s childbirth history. Interestingly, women who had given birth experienced less pain than those who had not.

These findings may support the use of simple pre-procedure questionnaire to identify risk factors, such as history of painful cervical screening or child-birth history, to identify patients at risk of severe pain during OPH. While more research is needed to confirm these results, this study highlights the importance of understanding individual risk factors to make OPH safer, less painful and more tolerable for all women.

## Introduction

Hysteroscopy is considered an indispensable procedure in diagnostic gynaecology(Paulo, Solheiro and Paulo, 2015). Historically, hysteroscopy required general anaesthetic, although modern hysteroscopes facilitate application to an outpatient pathway. This aligns with the NHS target that 75% of elective surgeries should be completed as outpatient procedures(Bailey *et al*., 2019; Marbaniang, 2019), associated with decreased completion times, no risk of general anaesthetic use and lower clinical costs(Bajaj *et al*., 2009; Darwin and Chung, 2013; Anderson, Walls and Canelo, 2017).

The transition from general anaesthetic demands a previously unrequired focus on pain management. Outpatient hysteroscopy(OPH) elicits pain in 85% of patients, with 15-34.8% experiencing severe pain(De Laco *et al*., 2000; Harrison *et al*., 2020; Mahmud, Smith and Clark, 2021). NICE guidelines recommend OPH as the primary diagnostic for common conditions such as heavy menstrual bleeding, endometrial pathology or dysmenorrhea(*NICE: Outpatient Hysteroscopy Quality statement 2* | *Heavy menstrual bleeding* | *NICE*, 2013, p. 2). As such, pre-COVID-19 estimates suggest as many as 71,000 OPH procedures are conducted in England annually(Elahmedawy and Snook, 2021). As financial demands prevent a reduction in outpatient procedures, understanding which women are at risk of severe pain stands as a critical domain for service improvement.

Preliminary audits identify a consistent spectrum of pain within OPH(De Laco *et al*., 2000; Cicinelli *et al*., 2007; De Carvalho Schettini *et al*., 2007; Harrison *et al*., 2020; Mahmud, Smith and Clark, 2021; Malu *et al*., 2023). The risk of pain is particularly problematic as strategies for perioperative pain management appear to be ineffective and insufficient(Ahmad *et al*., 2017). Only local anaesthetic reduces pain to a statistically significant extent (a reduction in pain intensity of 0.7/11), this reduction is “unlikely to be clinically significant”.

OPH has an estimated failure rate of 12%(Genovese *et al*., 2020), with pain frequently identified as the primary cause(Bettocchi *et al*., 2002; Guraslan *et al*., 2022). While OPH can be reattempted, it should not be ignored that this still exposes a patient to severe pain in the first instance. Additionally, most patients are likely to refuse a repeat attempt, putting themselves at risk of undiagnosed pathology or ongoing aversive symptomology(Genovese *et al*., 2020). Ideally, the initial failed outpatient procedure would be bypassed in the first instance. Optimising assessment maybe an option to identifying those at risk prior to the procedure. Variables such as anxiety, pain catastrophising, menopause status, chronic pelvic pain and previous childbirth experience are easily attainable, and may predict pain(Cicinelli *et al*., 2007; De Carvalho Schettini *et al*., 2007). Further investigation of quantifiable risk factors may help reduce exposure to unnecessary pain, and improve completion and satisfaction rates.

Hysteroscopy is an essential gynaecological diagnostic tool. The risk of severe pain presents a clear ethical and medical challenge. Predictively identifying patients at risk of pain, would facilitate an improvement in the quality of the service. This audit aims to investigate pre-procedural variables, and may stand to help stratify the patients at risk of severe pain and treatment failure in OPH.

## Method

### Sample

Audit data was collected, between 2009-2017 at the Royal Berkshire Hospital’s obstetrics and gynaecological department for the purposes of service evaluation. Data was collected from 804 patients who underwent OPH(M_age_=51.8y, s.d.=12.2y).

### Data Collection

Patients retrospectively rated their pain during their most recent cervical screening, and their last blood test sampling from the antecubital fossa, on an 11-point visual analogue scale(0-10). Following the procedure, patients completed an 11-item questionnaire about their OPH experience. Items included whether they would have OPH again and whether they’d recommend it to a friend/relative. Patients were asked if their comfort matched expectations, if they were satisfied with the experience, whether they were anxious before their appointment and whether the written information made them anxious. Patients also reported whether they have had previous OPH or hysteroscopy under general anaesthetic and if their procedure compared favourably to previous general anaesthetic hysteroscopies. Lastly, patients indicated whether they were given the opportunity to ask questions.

A second questionnaire was completed post-procedurally by the operating clinician. Clinical details relating to the procedure, findings and potential complications emerging from the OPH. How many ampules of local anaesthetic were applied during the procedure, alongside an estimate of the patient’s pain on a 5-point descriptive scale(none; discomfort; mild; moderate; severe), were provided.

To eliminate response bias, each questionnaire was completed in a double-blind procedure, with the patient and clinician unaware of the others’ responses.

### Clinical Procedure

All hysteroscopies included within the service evaluation were completed within an outpatient pathway. Patients were given a cervical block with up to 3 ampules of local anaesthetic(3% plain mepivacaine hydrochloride), applied based on the decision-making of the operating clinician. Most patients also required endometrial biopsy, wherein an endometrial sampler was used to collect a sample of tissue. Once the procedure was concluded, patients completed their post-procedural questionnaire. At this point, the operating clinician would complete their post-procedural report.

### Data Analysis

A-priori analyses focused on variables that may be identified presurgically. Therefore, variables which include a retrospective component, such as “was the procedure better/worse than expected” were not included. Therefore, the independent variables included were: biological(postmenopausal status); demographic and medical history(history of gynaecologic exams, number of births, previous pregnancy complications, previous birth modality); and procedural details(presurgical analgesia, cervix preparation, endometrial biopsy, hysteroscopy indication, anaesthetic used, number of polyps removed, hysteroscopy outcome & surgery performed). Preprocedural patient estimates of pain from their last cervical screening and blood test were also included.

Pre-processing was required to prepare data for stepwise logistic regression analyses. Pain ratings on an 11-point ordinal scale, including retrospective cervical screening/blood test ratings and OPH ratings themselves, were recoded to categorical format as follows: “no pain”(0), “mild”(1-3), “moderate”(4-6) and “severe”(7-10). Responses of “cannot recall”, and “not done” were retained.

All other variables remained in their original categorical format, as linearity could not confidently be assumed. Missing data were excluded from analyses, rather than applying interpolation.

Data analysis was performed using RStudio(Posit team, 2024) and the MASS package(Venables & Ripley, 2002).

### Ethics

All data was collected during routine medical treatment. In accordance with the NHS’s confidentiality code of practice, the anonymity of the patient and medical staff was preserved throughout the collection of data, its storage and analysis. The project was completed within the ethical framework of a service evaluation and clinical audit. Permission to analyse and disseminate the data was obtained via the local Research & Development department, in line with the National Research Ethics Service guidance on the completion of clinical audits

## Results

Of the initial 804 patients, after removing participants with missing data, the final dataset included 409 full datasets. All dependent variables were originally entered in an ordinal regression model, and then a stepwise model was selected based on the AIC index.

The resulting stepwise ordinal logistic regression was conducted to examine the relationship between various predictors and patient-reported pain levels, categorized as “no pain,” “mild,” “moderate,” and “severe” pain. An analysis of deviance(Type II ANOVA) was performed to confirm the presence of five variables significantly associated with intra-operative pain(Table 1): Number of births(0-5+), hysteroscopy indication(premenopausal bleeding/discharge, postmenopausal bleeding and IUD removal), hysteroscopy outcome(normal, atrophic, polyps, failed procedure or other), surgery performed(diagnostic with biopsy or polypectomy), and patient’s pain estimate score from their previous cervical screening(none, mild, moderate, severe, can’t recall or never had).

**Table 1.** Analysis of Deviance for Predictors of Patient Pain.

| Predictor | Likelihood Ratio<br>(Chi-square) | Degrees of<br>freedom | p-value |
| --- | --- | --- | --- |
| Number of Births | 16.02 | 5 | .006 ** |
| Hysteroscopy Indication | 5.38 | 2 | .06 |
| Hysteroscopy Outcome | 14.17 | 4 | .006 ** |
| Surgery Performed | 4.71 | 3 | .19 |
| Cervical Screening Pain | 46.66 | 6 | <.001 *** |

To investigate the nuances across each of the levels of the variables, the coefficients from the linear regression model are presented in Table 2. The directionality of the standardised beta(b) or z-value signifies if the variable is associated with higher(positive) or less(negative) pain. Within each variable(i.e. number of births), the statistics associated with each level represent the comparison to the lowest level within that variable. For example, 2 births are associated with less pain, in relation to 0 births.

**Table 2.**
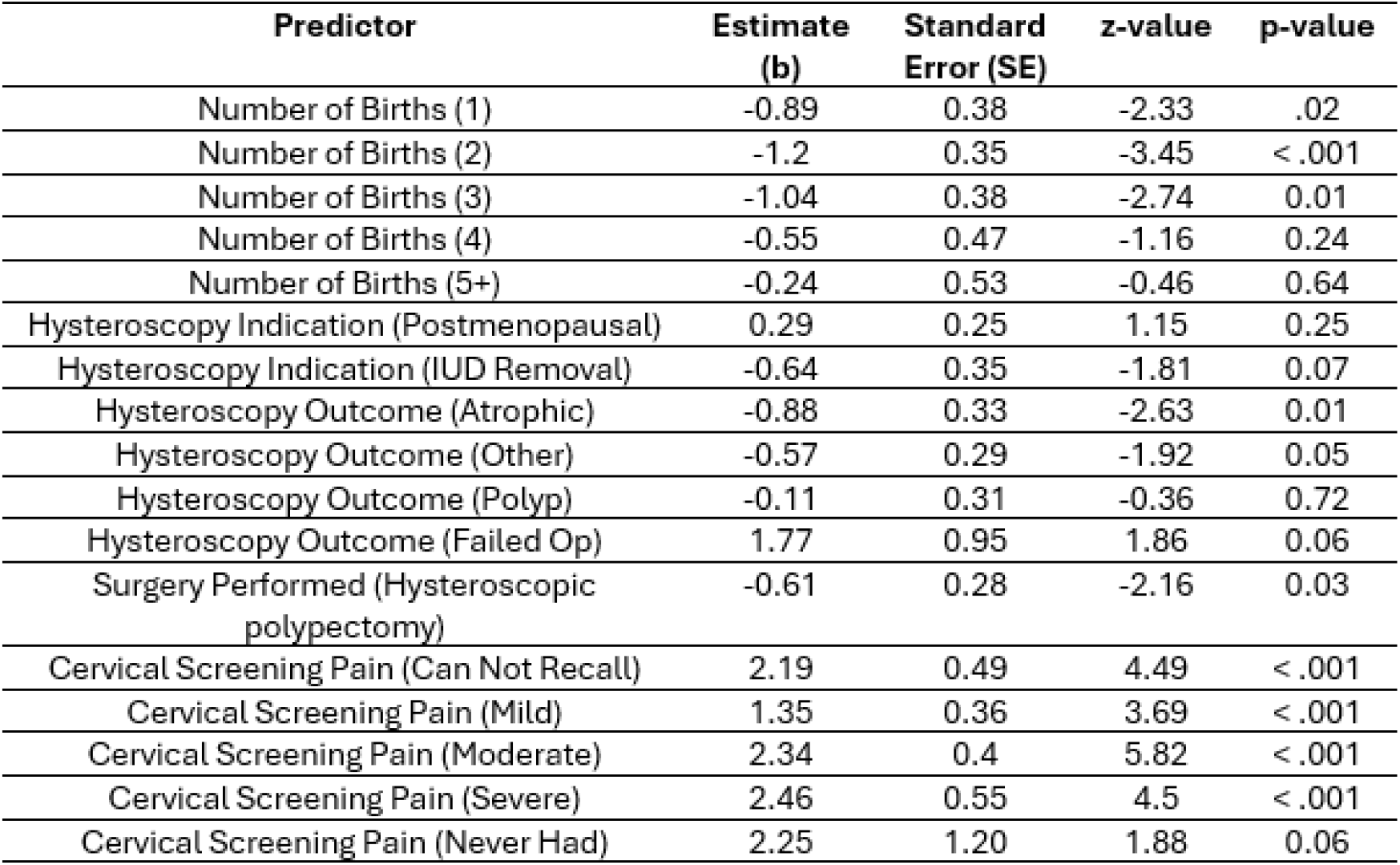
Ordinal Logistic Regression Coefficients for Predictors of Patient Pain(95% Confidence Intervals)

| Predictor | Estimate<br>(b) | Standard<br>Error (SE) | z-value | p-value |
| --- | --- | --- | --- | --- |
| Number of Births (1) | -0.89 | 0.38 | -2.33 | .02 |
| Number of Births (2) | -1.2 | 0.35 | -3.45 | < .001 |
| Number of Births (3) | -1.04 | 0.38 | -2.74 | 0.01 |
| Number of Births (4) | -0.55 | 0.47 | -1.16 | 0.24 |
| Number of Births (5+) | -0.24 | 0.53 | -0.46 | 0.64 |
| Hysteroscopy Indication (Postmenopausal) | 0.29 | 0.25 | 1.15 | 0.25 |
| Hysteroscopy Indication (IUD Removal) | -0.64 | 0.35 | -1.81 | 0.07 |
| Hysteroscopy Outcome (Atrophic) | -0.88 | 0.33 | -2.63 | 0.01 |
| Hysteroscopy Outcome (Other) | -0.57 | 0.29 | -1.92 | 0.05 |
| Hysteroscopy Outcome (Polyp) | -0.11 | 0.31 | -0.36 | 0.72 |
| Hysteroscopy Outcome (Failed Op) | 1.77 | 0.95 | 1.86 | 0.06 |
| Surgery Performed (Hysteroscopic polypectomy) | -0.61 | 0.28 | -2.16 | 0.03 |
| Cervical Screening Pain (Can Not Recall) | 2.19 | 0.49 | 4.49 | < .001 |
| Cervical Screening Pain (Mild) | 1.35 | 0.36 | 3.69 | < .001 |
| Cervical Screening Pain (Moderate) | 2.34 | 0.4 | 5.82 | < .001 |
| Cervical Screening Pain (Severe) | 2.46 | 0.55 | 4.5 | < .001 |
| Cervical Screening Pain (Never Had) | 2.25 | 1.20 | 1.88 | 0.06 |

A likelihood ratio test comparing the stepwise model to a null(intercept-only) model indicated that the predictors collectively explained a significant proportion of the variance in pain levels(χ2(20) = 78.96, p<.001). The Cox Snell Pseudo R-squared for the model was 0.18.

Estimates of previous cervical screening pain emerged as the strongest predictor of pain(χ2 = 46.66,p < .001). Compared to those who remembered no pain, patients who estimated mild cervical screening pain were more likely to report higher pain(b = 1.35,p < .001). Those who recalled moderate cervical pain had an even greater likelihood of reporting higher pain(b = 2.34,p < .001), with severe cervical screening pain presenting a further increase(b = 2.46,p <.001). These findings highlight the enduring impact of prior procedural pain on future experiences. Compared to those who recalled no pain, even patients who could not recall(b = 2.19,p < .001) or who had never experienced cervical screening(b = 2.25, p = .06) were more likely to report higher pain, which may reflect heightened anxiety or unfamiliarity with similar gynaecological procedures.

Number of births significantly predicted pain(χ2 = 16.02,p = .006), with results indicating that one(b = −0.89,p = 0.02), two(b = −1.20,p < 0.01), and three(b = −1.04,p < 0.01) births were significantly associated with lower pain, compared to no births. Patients with four or more showed no significant differences in pain(p > 0.05). This indicates a lack of linearity and variability in how childbirth history influences pain perception(Figure 2). As the number of births include a combination of Caesarean and vaginal deliveries, variability was investigated and found to be similar across the sample(Figure 1).

**Figure 1.**
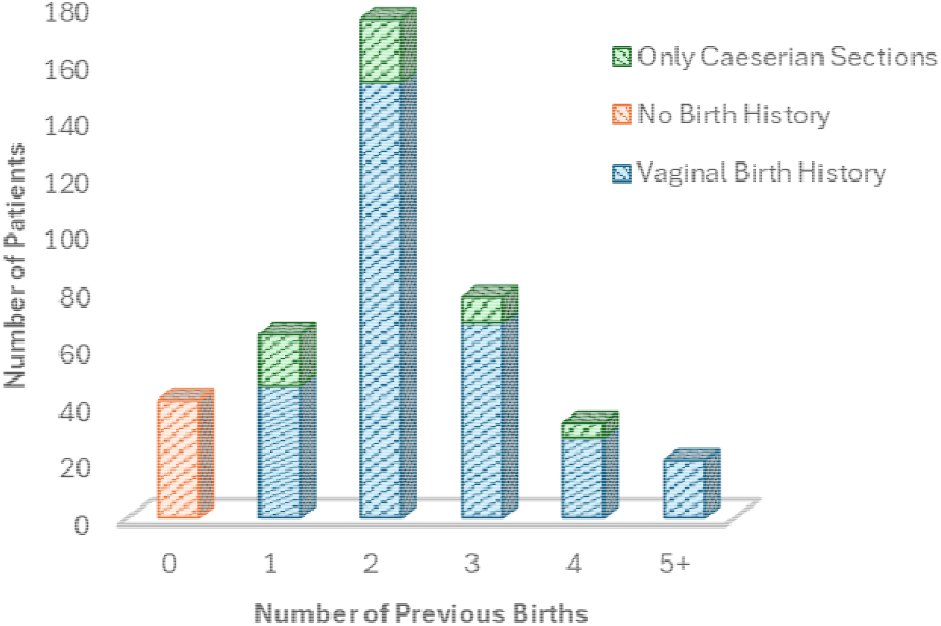
Representation of modes of delivery across number of births across sample.

**Figure 2.**
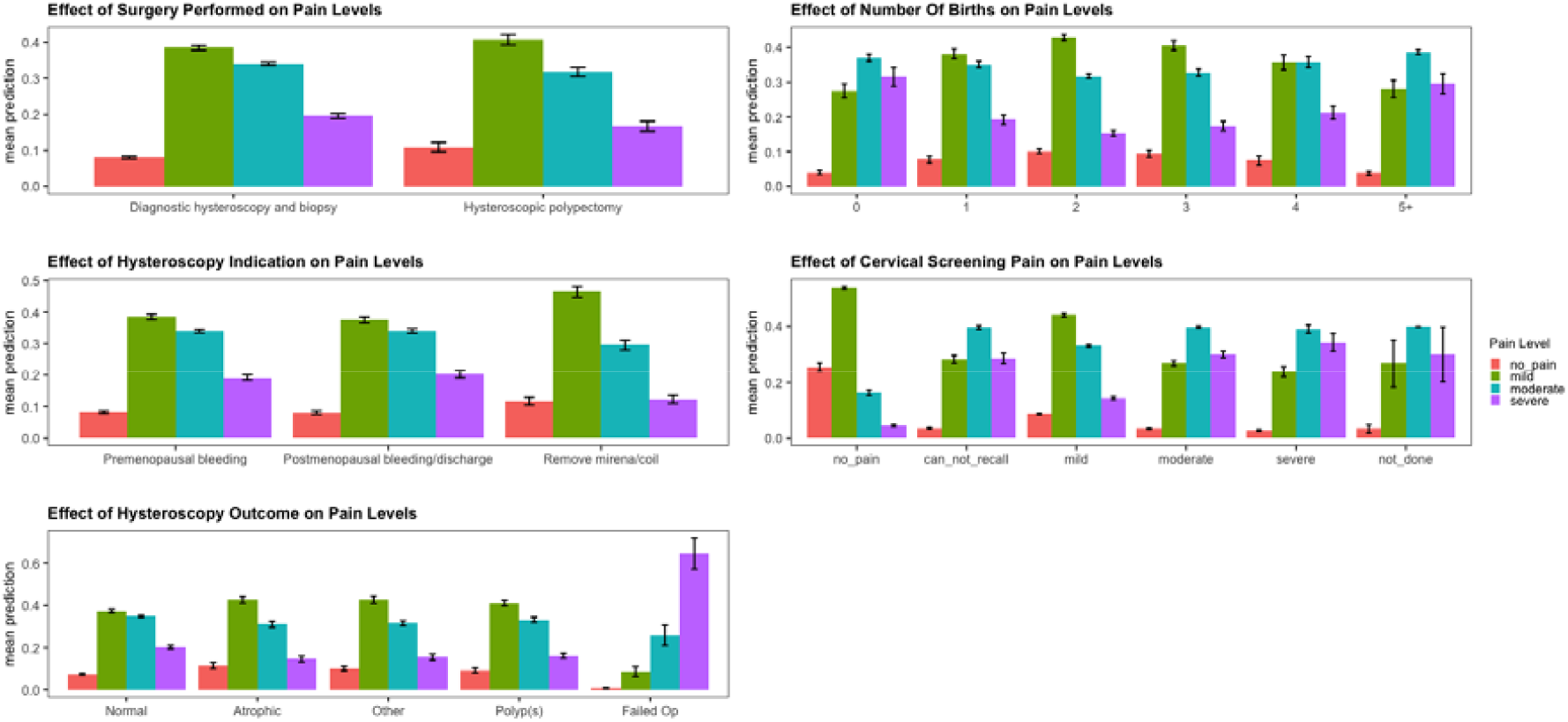
Visualisation of probabilities associated across the levels of the five significant variables identified as significant predictors of outpatient hysteroscopy pain, bars shown are standard error.

Although hysteroscopy indication was not a significant predictor(χ2 = 5.38,p = .06), regression coefficients indicated nuances in pain. While non-significant, IUD removal was somewhat associated with lower pain(b = −0.64,p = 0.07), than OPH for pre-menopausal bleeding. Patients undergoing hysteroscopy for postmenopausal bleeding or discharge showed no significant differences in pain intensity compared to other indications(p = 0.25).

The outcome of the procedure and pathological inferences were significantly associated with pain intensity(χ2 = 14.17,p = .006). Surprisingly, a diagnosis of atrophy was significantly associated with lower pain(b = −0.88,p = 0.01) than normal pathology. “Other” diagnoses, including fibroids, abnormally thickened endometrium(suspected hyperplasia) and scarring, were not significant, but tended towards lower pain(b = −0.57,p = 0.05). Collectively, these findings were not anticipated, although may be a feature of the larger variance within normal classifications, which comprised 51% of the sample, as opposed to atrophy(12%) or other(13%). Polyp diagnoses and failed operations did not show significant differences(p > 0.05), although the latter is most likely due to a low sample size after data pre-processing(n=6), with the incidence of severe pain appearing substantial within this subsample(Figure 2).

The regression coefficient for polypectomy was significant(b =-0.61,p = 0.03), indicating that patients undergoing this procedure were more likely to report lower pain levels compared to those undergoing diagnostic hysteroscopy, which included an endometrial biopsy.

### Thresholds for Pain Levels

To evaluate the overall risk of pain, cumulative probabilities of reporting no, mild, moderate or severe pain during the procedure were calculated(Table 3).

**Table 3.** Thresholds for Patient Pain Levels.

| Intercept | Estimate | Std. Error | t-value | p-value |
| --- | --- | --- | --- | --- |
| No Pain to Mild | -2.26 | 0.47 | -4.84 | <.001 *** |
| Mild to Moderate | 0.32 | 0.46 | 0.68 | .5 |
| Moderate to Severe | 2.1 | 0.47 | 4.49 | <.001 *** |

The intercepts represent the boundaries for cumulative probabilities of reporting pain at each level and determine the likelihood of reporting a higher level of pain instead of remaining in a lower level, considering all predictors in the model

The thresholds are presented in the log-odds form, but by using the formula below they were converted into probabilities using the formula:

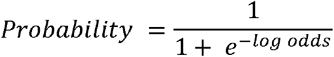

The threshold for the transition from “no pain” to “mild pain” corresponded to a 9.4% probability of reporting “no pain.” This represents an 90.6% probability of a patient reporting at least “mild pain,” during OPH. Primarily, this suggests that “no pain” is relatively uncommon unless specific predictors substantially increase the odds. The threshold for the transition from “mild pain” to “moderate pain” corresponded to a cumulative probability of 42.1% to report “moderate pain” or higher. However, this transition was not statistically significant(p =0.5), suggesting weak differentiation between these two categories. The threshold for the transition from “moderate pain” to “severe pain” indicated a 10.9% probability of reporting “severe pain”. This reduction is statistically significant(p<.001), highlighting that “severe pain” is relatively uncommon and typically requires the presence of specific predictors. Ultimately, these odds indicate hysteroscopy is predominantly likely to elicit mild-to-moderate pain.

## Discussion

Outpatient hysteroscopy(OPH) is frequently accompanied by significant pain(De Laco *et al*., 2000; Harrison *et al*., 2020; Mahmud, Smith and Clark, 2021). This highlights an urgent need for a deeper understanding of risk factors contributing to pain during OPH. Data was analysed from 409 patients and the results highlighted the importance of two types of pre-procedural variables in estimating pain. *Procedural* details, such as type of required surgery, pathological diagnosis and clinical indication. *Presurgical* details pertaining to the patient themselves, such as the retrospective estimate of pain for a previous cervical screening or history of childbirth. Importantly, this data is easily acquired and could be a potentially pragmatic step for identifying patients who are most at risk of experiencing a painful OPH.

National service and budget pressures necessitate the transition of onerous inpatient procedures to an outpatient setting(Marbaniang, 2019), but cost-saving benefits must be considered alongside patient safety and well-being. Pain is common within outpatient procedures(Vadivelu *et al*., 2016), and a primary cause of treatment failure(Bettocchi *et al*., 2002; Genovese *et al*., 2020). The risk of chronic postsurgical pain(CPSP) in outpatient procedures is equivalent to inpatient settings(Althaus *et al*., 2012; Gerbershagen *et al*., 2013) and peri-procedural pain intensity is robustly correlated with the risk of developing CPSP(Katz, Weinrib and Clarke, 2019). Multiple trials and audits have quantified the landscape of acute pain during outpatient hysteroscopy(De Laco *et al*., 2000; Genovese *et al*., 2020; Harrison *et al*., 2020; Mahmud, Smith and Clark, 2021), highlighting a need for OPH pathway optimisation.

Retrospective estimates of pain during cervical screening were the strongest predictor of OPH pain. Cervical screening is a common gynaecological procedure for women between the ages of 25-64(National Health Service, 2023), wherein pain is a possible consequence, with an incidence risk between 20-24%(Crombie *et al*., 1995; Hoyo *et al*., 2005) and average pain intensity estimated between 2-5 out of 10(Simavli *et al*., 2014; Yen *et al*., 2023). Uptake for cervical screening is high(National Health Service, 2023), presenting an opportunity for a presurgical variable that most OPH patients would be familiar with. Inpatient pain sensitivity assessment, predominantly via the Pain Sensitivity Questionnaire(PSQ; Ruscheweyh et al., 2009) is a promising approach, and can predict postsurgical clinical outcomes, central sensitisation(Tuna *et al*., 2018) and higher analgesia uptake. Notably, retrospective ratings of previous blood test pain did not predict OPH pain, suggesting the domain-specificity of cervical screening may be valuable.

Experience of childbirth was also associated with lower pain, regardless of whether via vaginal or Caesarean delivery. There is a paucity of literature regarding the long-term alterations in pain processing that follow childbirth. Experimentally, there is limited evidence that suggests nulliparous women have lower pain thresholds than parous women(Hapidou and DeCatanzaro, 1992), supporting anecdotal evidence that “nothing compares to childbirth”. Nulliparous patients are also more likely to experience more pain during early labour and during IUD fitting(Lowe, 2002; Rahman *et al*., 2024), corroborating our findings of sensitivity to gynaecological procedural pain. A limitation of this finding is that the dataset failed to discriminate between elective and emergency c-sections, which could plausibly have implications for pain sensitivity and limits the interpretability of this finding.

IUD removal was associated with a lower risk of OPH pain. While the gynaecological management of IUDs can frequently be associated with pain(Callahan *et al*., 2019), this is mostly associated with IUD insertion. The extraction of IUDs via OPH take less than 5-minutes(Asto and Habana, 2018), and requires no endometrial biopsy, unlike diagnostic inspections for pre/post-menopausal bleeding. Unexpectedly, polypectomies were also associated with a lower risk of pain than diagnostic hysteroscopy. Again, this potentially reflects the risk of pain with endometrial biopsy, which was included within nearly all diagnostic hysteroscopies. But this may also represent a limitation, wherein data failed to specify if polypectomies were cervical(requiring no entry to the uterus) or endometrial, with the former likely being less painful. Similarly, the size of the polyp has implications for pain, with large polyps frequently requiring endometrial biopsy(Spadoto-Dias *et al*., 2016), and associated with more intense pain(Litta *et al*., 2008). Collectively, these findings likely highlight to risk of pain with endometrial biopsy, as well as identifying the need for greater specificity in the evaluation of pain during polypectomy.

Lastly, while failed procedures were not significant within the model, visualisation of the data(Figure 2) may indicate a risk of severe pain is possible, however this should be interpreted cautiously due to low sample within this group, with only 29 failed operations across the entire sample(failure rate=3.87%).

Future directions require further empirical investigation of the predictive capabilities of the identified variables. Importantly, even variables associated with a lower risk of pain can still elicit painful OPH and should not be neglected or ignored. Sedation doesn’t appear to reduce post-operative pain ratings following colonoscopy(Seip *et al*., 2010) and pharmacological reductions in pain are unlikely to be clinical-meaningful(Ahmad *et al*., 2017). This suggests more liberal pain management alone is unlikely to be an effective optimisation and highlights the value of stratified assessment. Acute pain sensitivity is malleable to psychological interventions, such as brief mindfulness-based interventions (Reiner *et al*., 2016) or virtual reality-based distraction(Malloy and Milling, 2010). However, there are substantial barriers to applying these interventions to a procedure with an average completion time of around 5-minutes in a busy, clinical environment(Cicinelli *et al*., 2003). The results of this service evaluation suggest that easily acquired presurgical variables may be valuable for highlighting patients who are at risk of pain during their procedure.

## Data Availability

The participants of this study did not give written consent for their data to be shared publicly, so due to the sensitive nature of the research supporting data is not available.

## Acknowledgements & Declaration of Interest

The authors report there are no competing interests to declare

